# The COVID-19 Consequences of College Class Continuity Calculator: A Tool to Provide Students and Administrators with Estimated Risks of Returning to Campus

**DOI:** 10.1101/2020.07.31.20165761

**Authors:** Sharon Bewick, Erika Ludden, Suzanne Robertson, Jeffrey Demers

## Abstract

As schools prepare for the start of the Fall 2020 semester, many are struggling to make decisions regarding whether or not to return to on-campus classes or whether to remain fully online. Unfortunately, there is no ‘one-size-fits-all’ answer, and schools must balance their own risks against the costs of remote learning. We present a tool that integrates information about study body composition with predictions of COVID-19 infection rates in order to provide clarity and insight into the decisions facing colleges and universities nationwide. Our tool is freely available and currently hosted at the following location: https://bewicklab.shinyapps.io/covid-1/

## Introduction

Beginning in March 2020 with the detection of early COVID-19^1,2^ outbreaks in the U.S.^3^, teaching institutions countrywide quickly shifted to remote online classes^4-7^. At the time, there was a sense that this was a temporary stopgap measure – that students would return to campus to resume face-to-face (f2f) classes in a few weeks, months or, at the very least, by the start of the Fall 2020 semester. Since March, however, the spread of COVID-19 within the U.S. has gotten worse. This has necessitated a variety of heavy-handed measures, including stay at home orders, self-quarantine, and implementing legislature that promotes social distancing to limit disease spread. Despite this, cases of COVID-19 continue to grow, particularly in regions in the U.S. South and Mid-West. Indeed, within the last several weeks, there have even been renewed calls from experts urging the country to shut down once again in order to get control of the epidemic^8^.

The continued impact of COVID-19 on public health across the country creates a high level of uncertainty and debate over whether or not colleges and universities should resume in-person classes for the Fall 2020 semester. At a minimum, continuity plans for the fall semester must factor in student safety, educational impact, accessibility, finances, and potential legal issues. On the financial side, for example, institutes of higher education have already endured unprecedented losses as a result of the transition to online learning in March. Compounding those losses with lost revenue due to lower enrollment, a lack of on-campus housing, and potentially reduced tuition for online classes would be devastating, and would result in lay-offs, furloughs and possibly even the permanent closure of certain schools. In terms of educational costs, it is clear that certain experiences – for example studio or lab classes – are more difficult in an on-line format. And even for courses that can transition online relatively easily, issues of equity remain. For example, lack of access to high quality internet is likely to disproportionately impact marginalized populations, and this must be addressed to make college courses fair, equitable and accessible to all.

While both financial and educational arguments suggest that a return to f2f classes for Fall 2020 is optimal, the health and safety risks of doing so are extreme. Returning students to campus would involve an unprecedented, mass, multistate migration across the country. This would introduce COVID-19 from regional ‘hot-spots’ to locations with lower levels of spread. It would also make any efforts to contact-trace almost impossible. Not only will this have the effect of worsening the outbreak in regions where the virus has, so far, been successfully contained, but also, it will pose a risk for students, faculty and staff and even the broader members of local communities in towns and cities where institutes of higher education exist. These health risks bring financial concerns of their own. If, for example, faculty, staff or students suffer severe morbidity or mortality as a result of an on-campus outbreak, lawsuits may be inescapable. As move-in dates quickly approach in the following weeks, both students and administrators need to understand the risks of returning to campus to make informed decisions going forward.

One of the challenges faced by students, colleges and universities in making the decision to stay open or to move to remote learning is that all institutes of higher learning are unique. They draw different numbers of students from different locations across the U.S. and, as a consequence, the risks of one school opening for f2f classes are not necessarily the same as the risks of another school opening. In order to provide students and administrators with a more individualized assessment of the risks of student return to campus, our team has developed the ‘COVID-19 Consequences of College Continuity Calculator’. This tool provides estimates of the number of students expected to return to campus carrying COVID-19, as well as the states from which those students are expected to arrive. Specifically, our tool combines state-level demographic data from each college or university with state-level predictions from Institute for Health Metrics and Evaluation (IHME) models to determine COVID-19 infection rates in returning students. Our tool has the ability to generate predictions unique to the geographical representativeness of 1440 school’s student bodies. These predictions provide an understanding of the potential risks unique to individual universities based on student body size, geographic location, and school opening strategy. Our hope is that this tool will provide a missing informational component that can be used to factor into the informed decision making of both administrators and students in returning to campus.

The online tool ‘COVID-19 Consequences of College Class Continuity Calculator’ is currently available at the website: https://bewicklab.shinyapps.io/covid-1/

## Methods

All code is available at the following Github repository: (https://github.com/bewicklab/COVID19_Consequences_of_College_Class_Continuity_Calculator)

### Student enrollment data

Student enrollment data was obtained using the custom Python code ‘class_names.py’ to webscrape data on college enrollment from the College Factual website. Briefly, we collected a list 1440 of colleges and universities from: https://www.collegefactual.com/search/.

For each school, we then used this list to pull the relative counts of the number of students from each state from the following set of websites: https://www.collegefactual.com/colleges/university-name/student-life/diversity/chart-geographic-diversity.html

Next we rescaled these relative counts such that the total count summed to the population of the full undergraduate student body at each school. Total undergraduate population size was determined from the following set of websites: https://www.collegefactual.com/colleges/university-name

Both total student body size and state-by-state breakdowns of the student body are based on enrollment during the 2017-2018 academic year, since this was the most recent data available. Predictions are less likely to be accurate for any school that has experienced a significant change in enrollment in the last 1-2 years, or schools with enrollment significantly affected by COVID-19.

### COVID-19 infection rates

Predictions for the number of COVID-19 infections in each state are based on IHME models downloaded from the IHME: COVID-19 Projections website at (http://www.healthdata.org/covid/data-downloads). These models provide lower estimates, upper estimates and mean estimates of the number of daily deaths in each state. To convert from daily deaths to new infections, we first assumed that it takes 19 days between becoming infectious (symptomatic or a positive polymerase chain reaction (PCR) test, hereafter referred to as an ‘active’ infection) and dying. Thus, we assumed that all deaths on date *t* were due to new active infections on date *t* – 19. We then used the infection fatality ratio (IFR) to convert from daily active infections that resulted in death to total daily active infections. We did this by dividing the number of deaths 19 days later by the IFR. For the IFR, we used the current best estimate of IFR = 0.0065^9^.

Finally, to convert from new active infections to infection rate, we divided state-level predictions of active infections by the population size of each state (see equation (1)). State populations were obtained from the following website (accessed on 07/21/2020): https://simple.wikipedia.org/wiki/List_of_U.S._states_by_population

We used IHME mean estimates to provide our expected values, and IHME low and high estimates to provide ranges.

Although we anticipate updating our online tool weekly to account for changes in IHME estimates, all analysis in the current paper was based on data downloaded from the IHME: COVID-19 Projection website on 07/30/2020, and reflects IHME predictions updated on 07/14/2020. We did not take the most recent dataset, because lower bound estimates were missing. However, estimates for the state of New Hampshire were missing in the predictions for 07/14/2020; thus for New Hampshire, we used predictions from 07/22/2020 (which lack a lower bound). The custom Python code ‘IHME_predictions_just_states_by_death.py’ was used to identify state-level IHME predictions from downloaded datasets and to convert these to daily new active infections.

In our online tool, users are allowed to select between three different IHME models: ‘Mandates Easing’, ‘Current Projection’ and ‘Universal Masks’. Details regarding each of these models can be found on the IHME: COVID-19 Projections website. Briefly, ‘Mandates Easing’ assumes continued easing of social distancing mandates and that these are not re-imposed. ‘Current Projection’ assumes mandates are re-imposed for 6 weeks whenever daily deaths rates reach 8 per million. Finally, ‘Universal Masks’ assumes 95% mask use in public in every location, along with re-imposing social distancing mandates when daily deaths rates reach 8 per million.

### Probability that at least one student will return carrying COVID-19

The graphical user interface (GUI) for our online tool allows users to choose their school, the IHME model scenario, *m*, that they would like to use, the expected date for student return to campus, *d*, and estimates for both the incubation period (time between exposure and becoming infectious), *T*_*E*_, and the infectious period (time between becoming infectious and recovery or death), *T*_*1*_, of COVID-19. From this information, we first calculate the predicted rate of infection (both active infections and infections still in the incubation phase) in each state on date *d, P*_*d,S*_ as follows:

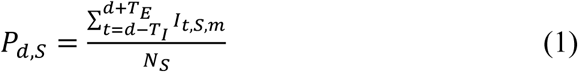

where *I*_*t,S,m*_is the number of new active infections predicted by IHME model *m* on date *t* in state *S*, and *N*_*S*_ is the total population size of state *S*. We note the sum in equation (1) is from *t* = *d−T*_*1*_ to *t* = *d*+*T*_*E*_. That is, we count all of the people who developed the disease up to *T*_*1*_ days *prior* to returning to campus, since these people are expected to still be infectious when they return. Likewise, we count all of the people who *will* develop the disease up to *T*_*E*_ days *after* returning to campus, since these people will have been exposed to COVID-19 in their home states prior to returning to campus. Using equation (1), we then calculate the probability of at least one student arriving on campus carrying COVID-19, *P*_*X*_, as follows:

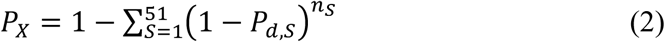

where *n*_*S*_is the number of students returning from state *S*, and the sum is over all 50 states, as well as the District of Columbia.

### Expected number of students returning with a COVID-19 infection

We calculate the expected number of students returning with an active COVID-19 infection, *E*_*n*_, as follows:

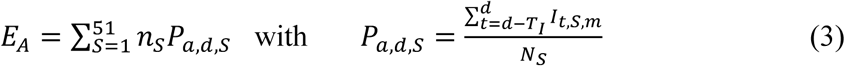

where *P*_*a,d,S*_ is the probability of a student having become infectious at any time point up to *T*_*1*_ days (infectious period) prior to arriving on campus. Similarly, we calculate the expected number of students returning to campus who are still in the incubation phase of a COVID-19 infection, *E*_*P*_, as follows:

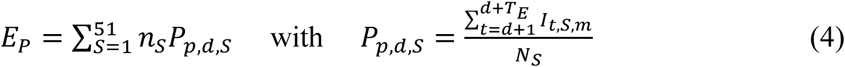

where *P*_*a,d,S*_ is the probability of a student becoming infectious at any time point from the day after arrival on campus up to *T*_*E*_ days later (incubation period). The total expected number of students that will arrive on campus carrying the COVID-19 virus, *E*_*C*_, is then given by:

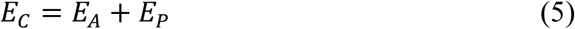

State-level predictions are given by the individual terms in equations (3) and (4), with their sum giving the state-level equivalents to equation (5).

### Effectiveness of proposed quarantine and testing strategies

A number of schools are proposing some combination of quarantine and testing in order to mitigate the introduction of COVID-19 on campus. To help assess the effects of these actions on the risk of COVID-19 during return to campus, our tool includes a prediction of the number of student infections that will be missed based on a school’s selected quarantine and testing protocol. Specifically, our GUI allows users to select the mandatory quarantine length, *Q*, the percentage of students that will be tested, *θ*, and the estimated false-negative rate, *ρ*, for their chosen testing method. The expected number of COVID-19 cases that will be missed is then be given by:

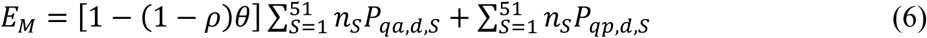

with 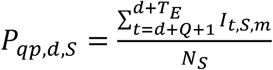 if *T*_*E*_. > *Q* and *P*_*qp,d,S*_ =0 otherwise, and 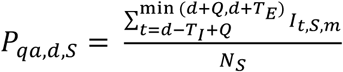. Here *P*_*qp,d,S*_is the probability of a student returning exposed and becoming infectious after the quarantine period is completed and testing has been administered, and *P*_*qa,d,S*_ is the probability of a student having an active Covid-19 infection at the time of testing. Equation (6) assumes that testing is conducted at the end of the quarantine period, and 100% of COVID-19 infections still in the incubation phase go undetected by available tests. In addition, a fraction of active COVID-19 infections also go undetected, based on the percentage of students tested, as well as the number of tests that give false-negatives, and thus fail to detect active cases.

### Parameters and model predictions

Unless otherwise noted, we assume an infectious period of 10 days, in keeping with recent Center for Disease Control (CDC) recommendations^10^. We also assume an incubation period of 5 days^11^ and a return to campus date of 08/15/2020. In what follows, we use these assumptions to explore the consequences of on-campus return across all 1440 institutes of higher education in our database. Specifically, we examine the number of students returning as COVID-19 carriers, both in absolute terms and in terms of rates of infection across the student body. We then examine how these outcomes depend on student body size, geographic location, and school opening date.

## Results

We begin by exploring trends across all colleges and universities in our database. For the worst-case scenario (‘Mandates Easing’), we find a mean(median) of 33.50(9) students returning to campus as COVID-19 carriers. This includes a mean(median) of 22.51(6) students returning with active cases, and a mean(median) of 10.99(3) students returning still in the incubation phase. In terms of incidence rates, we find a mean(median) of 4.8(3.5) cases per 1000 students, with 3.2(2.3) of these cases being active and 1.6(1.1) of these cases being in the incubation phase. Overall, the mean(median) probability of at least one student returning to campus infected with the virus is 96.14% (99.99%). Results for the other two IHME scenarios are shown in Table 1.

**Table 1:** Summary statistics across all colleges and universities for each of the IHME models and assuming *T*_*I*_ = 10 days, *T*_*E*_ = 5 days, and *d* = 08/15/2020

|  | Mandates Easing | Current Projection | Universal Masks |
| --- | --- | --- | --- |
| Mean probability of at least one student returning with COVID-19 | 96.14% | 96.07% | 86.12% |
| Median probability of at least one student returning with COVID-19 | 99.99% | 99.99% | 96.64% |
| Absolute Number of Students |  |  |  |
| Mean number of students returning with COVID-19 | 33.50 | 32.18 | 12.57 |
| Median number of students returning with COVID-19 | 9 | 9 | 3 |
| Mean number of students returning with an active case | 22.51 | 22.23 | 8.86 |
| Median number of students returning with an active case | 6 | 6 | 2 |
| Mean number of students returning in incubation phase | 10.99 | 9.95 | 3.71 |
| Median number of students returning in incubation phase | 3 | 3 | 1 |
| Per Capita Number of Students |  |  |  |
| Mean number of students returning with COVID-19 | 0.0048 | 0.0046 | 0.0017 |
| Median number of students returning with COVID-19 | 0.0035 | 0.0034 | 0.0013 |
| Mean number of students returning with an active case | 0.0032 | 0.0032 | 0.0012 |
| Median number of students returning with an active case | 0.0023 | 0.0023 | 0.00096 |
| Mean number of students returning in incubation phase | 0.0016 | 0.0014 | 0.00047 |
| Median number of students returning in incubation phase | 0.0011 | 0.0011 | 0.00039 |

For all three scenarios, the distribution of cases across schools was highly skewed, with most schools having <10 COVID-19 infections among the returning students, while a few schools had hundreds to more than 1000 expected cases (see Fig 1a, see also SI Fig. S1). This was at least partly driven by the wide range of school sizes, with larger schools having significantly more cases than smaller schools (Fig 1b and c, see also Fig. S1).

**Fig 1.**
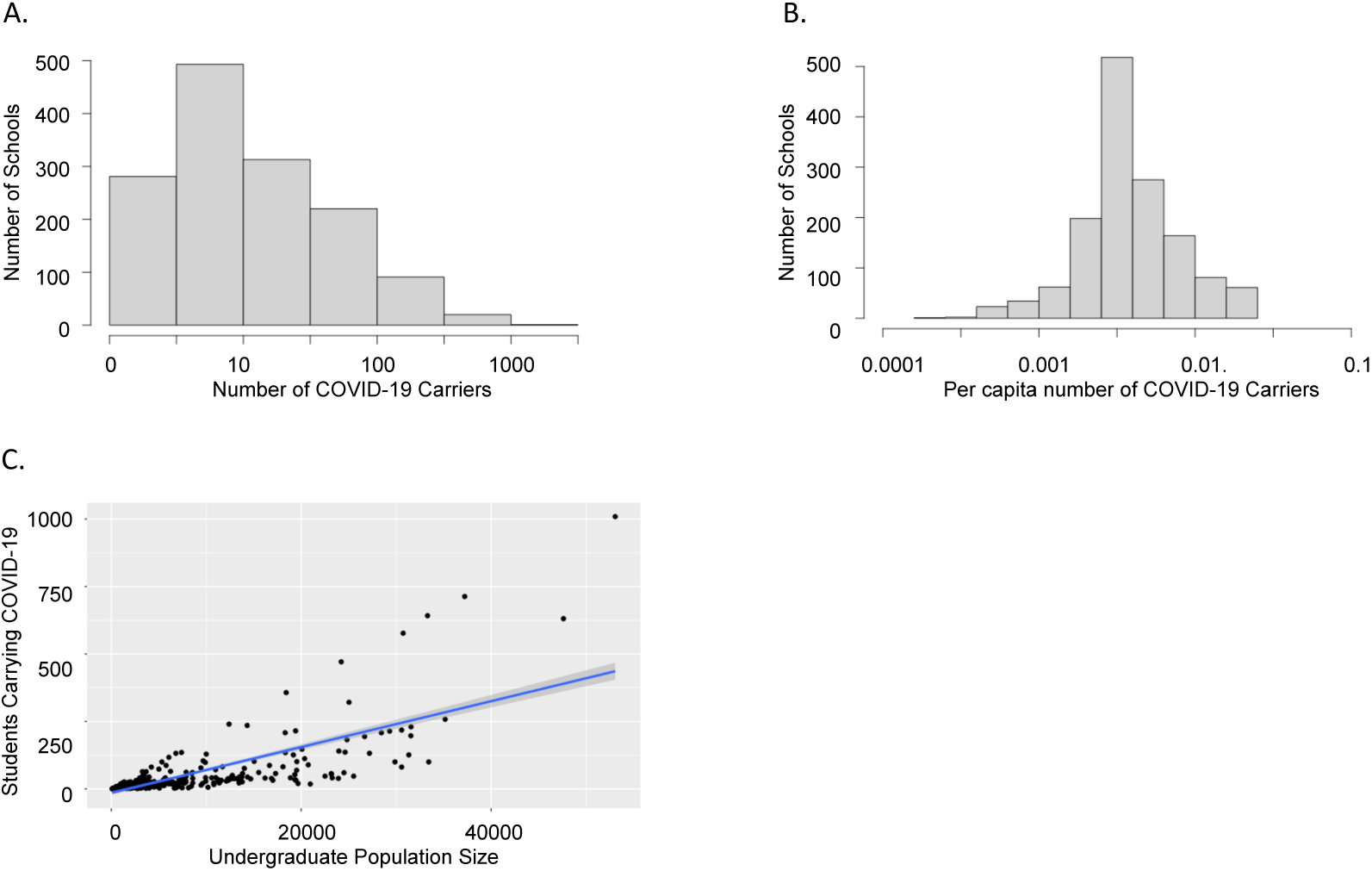
Summary of COVID-19 trends across all colleges and universities in our database. (A) Histogram showing the number of schools with different absolute numbers of students arriving on campus carrying COVID-19 (infected + incubating); (B) Histogram showing the number of schools with different per capita numbers of students arriving on campus carrying COVID-19; (C) Regression showing the relationship between school size and the absolute number of COVID-19 carriers expected to return to campus. Results are shown for the ‘Mandates Easing’ scenario

Another important factor driving differences in COVID-19 infections across schools was school location. Even though colleges and universities draw students from across the country, higher proportions of students frequently come from locations within state, and this is particularly true of state schools. Thus, we considered absolute numbers of infections and infection incidence rates among returning students as a function of state (see Fig 2, see also SI Fig. S2). Not surprisingly, states in ‘hot-spots’, for example Texas and Florida had higher numbers of students returning as COVID-19 carriers as compared to states with less severe outbreaks, for example West Virginia.

**Fig 2.**
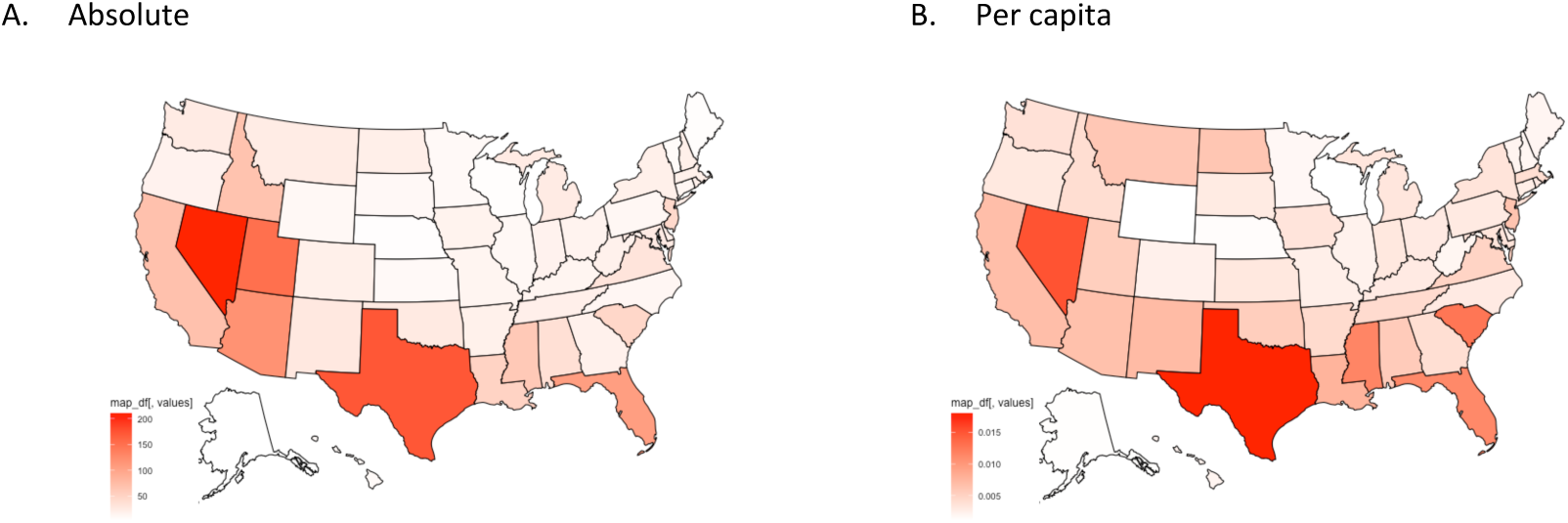
Summary of average (by state) COVID-19 trends across all colleges and universities in our database. (A) Absolute number of students arriving on campus carrying the COVID-19 virus; (B) Per capita number of students arriving on campus carrying the COVID-19 virus. Results are shown for the ‘Mandates Easing’ scenario.

One of the risks of on-campus classes in Fall 2020 is the mass migration of students across the country. This will disperse students from hot-spot regions to regions with controlled or mostly controlled outbreaks, exacerbating efforts at disease containment. For this reason, we also considered the states most at risk of introductions from other locations. This is a function of both the degree to which schools within a state draw from out-of-state student populations, as well as the level of outbreak in-state. This is shown in Fig. 3. As expected, states like West Virginia and Alaska that are not currently seeing high levels of COVID-19 infection are negatively impacted by the decision to move students back to campus. Meanwhile, states like Texas are not impacted at all – indeed, Texas is responsible for many of the imported cases to other states.

**Fig 3.**
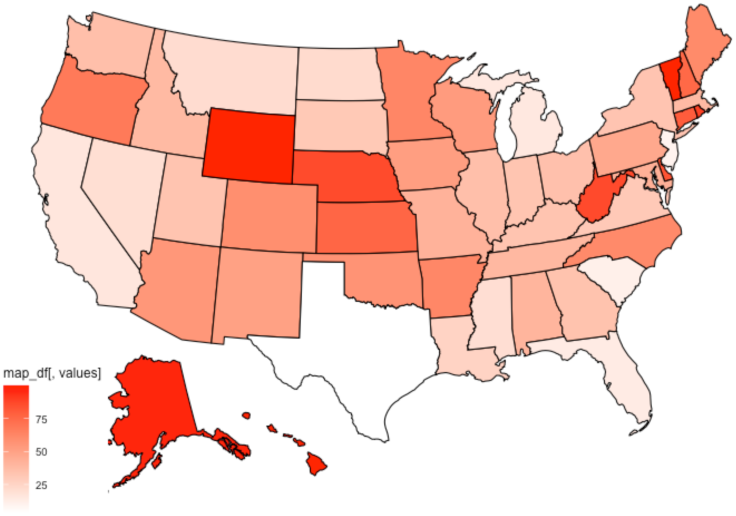
Summary of average (by state) percentage of COVID-19 cases that are from out-of-state students. Results are shown for the ‘Mandates Easing’ scenario.

In order to mitigate the risk of f2f classes in Fall 2020, many schools are contemplating a delayed start. Therefore, we considered the impact that this would have on infection rates among returning students. This is shown in Fig. 4 for all three IHME scenarios.

**Fig 4.**
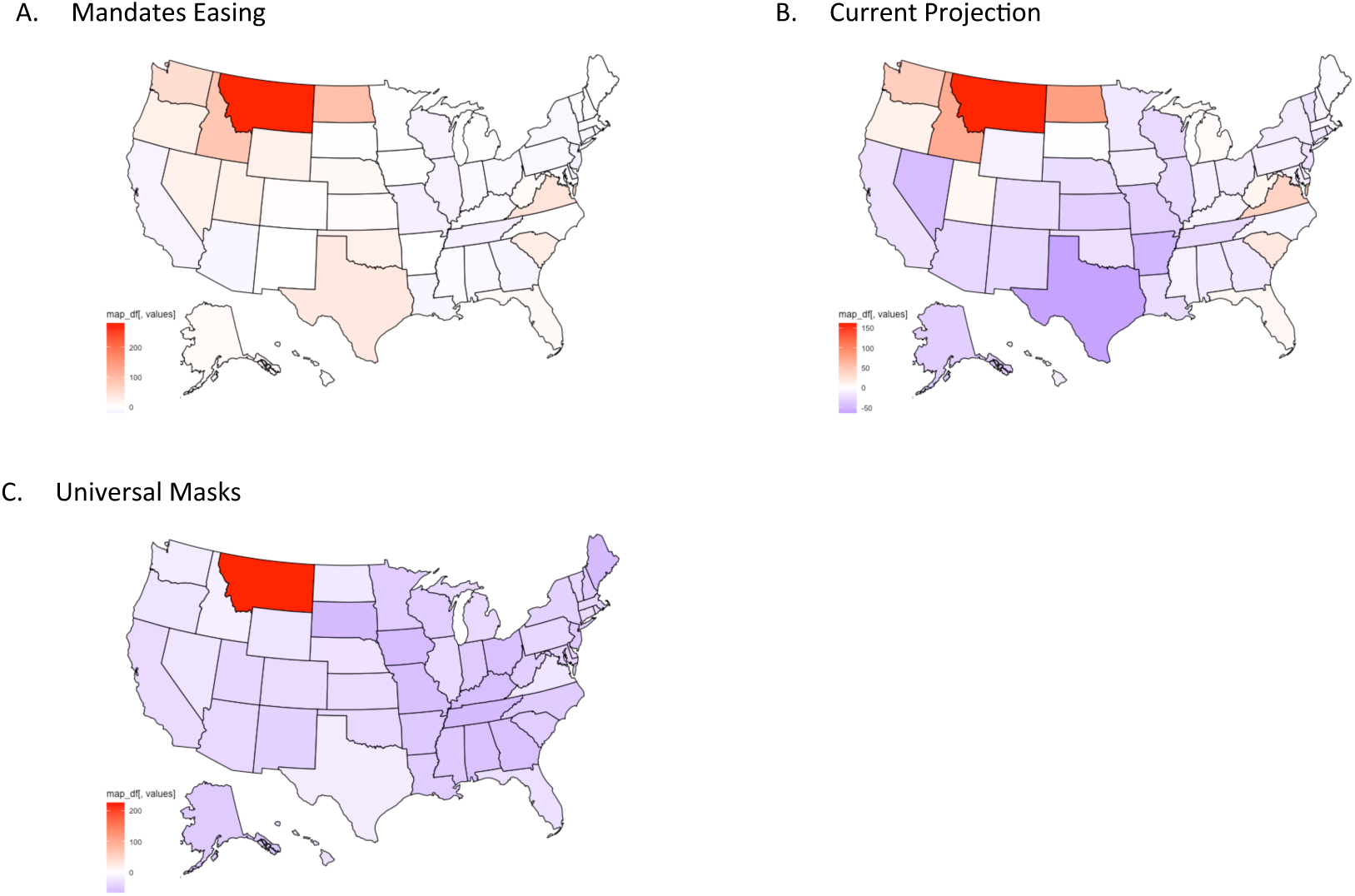
Summary of average (by state) COVID-19 temporal trends, showing the effects of delaying classes by 5 weeks. Values are reported as percent changes in the number of students arriving carrying the COVID-19 virus for the three different IHME scenarios: (A) Mandates Easing, (B) Current Projection and (C) Universal Masks

Here we assume a five week delay, consistent with current planning at Clemson University, where classes have been delayed from an original 08/17/2020 start date to an 09/21/2020 start date. Notice that states are colored by percent change – thus, for example, Montana has a large increase in all three maps. This is not because Montana has a particularly large increase in absolute numbers of infected students over the delay period, but rather, because infection rates on 08/17/2020 are very low, such that even modest increases in infection rates give dramatic percent changes. Clearly, whether or not a delay is helpful is strongly contingent on how public health measures are implemented. For the ‘Mandates Easing’ scenario, there was an overall 10.15% increase in the rate of infection in returning students (averaged across states; there was a 1.70% increase when averaged directly across schools), suggesting that a five week delay would not be beneficial and, if anything, could be detrimental. However, in the ‘Current Projection’ scenario, there was a 6.54% decrease (averaged across states; there was an 11.57% decrease when averaged directly across schools) in the rate of infection, providing hope that a delay in return to campus could be beneficial. Even more encouragingly, in the ‘Universal Masks’ scenario, there was a 36.53% decrease (when averaged across states; there was a 40.51% decrease when averaged directly across schools) in the rate of infection. Thus a delayed start to on-campus classes could be very effective if mask adoption can be successfully implemented nationwide.

## Discussion

We have developed an online tool that can help both students and administrators estimate the level of risk of returning to f2f classes for Fall 2020. For any given school, our tool predicts the number of returning students who will be carrying COVID-19. This estimate is broken down into students who have an active infection, as well as students who are in the incubation phase. The distinction between active infection and incubation phase is important, since only active infections are likely to be detected by a polymerase chain reaction (PCR) test for the COVID-19 virus. Thus, without any mandatory quarantines, even testing 100% of students, and assuming a testing false negative rate of 0%, students still incubating the virus will be allowed into the community with the ability to spread COVID-19.

To account for mitigation strategies, our tool enables users to select quarantine and testing combinations to see how this will impact the risk of re-opening campuses. As suggested above, we assume that all infections that are in the incubation phase evade detection. To prevent these infections from being missed, mandatory quarantine is necessary. For active infections, we assume that a certain number are missed as a result of incomplete testing of the student body, as well as imperfect testing results. Again, the goal of this component of our tool is to help administrators decide on the best balance of risks versus financial considerations based on their testing capacity, ability to quarantine students, etc.

While we find very few schools that have little to no risk of COVID-19 infections, we do find that most schools will have <10 imported cases. There are, however, other schools with large numbers of infections in the returning student body. Not surprisingly, most of these schools are located in ‘hot-spot’ states. For example, under the ‘Mandates Easing’ scenario, eight out of ten schools with the highest number of students returning infected are in Texas, while the other two are in Florida (1. Texas A&M University – College Station (1009), 2. University of Central Florida (744), 3. The University of Texas at Austin (743), 4. University of Houston (713), 5. Texas State University (662), 6. The University of Texas at Arlington (642), 7. Florida International University (631), 8. University of North Texas (598), 9. Texas Tech University (577), 10. The University of Texas at San Antonio (510)). These are also primarily state schools, with large student bodies. Whether or not re-opening is a viable strategy for these types of schools remains an open question, and probably depends on the capacity of these schools to test and quarantine incoming students.

Although our tool is useful for helping to guide decision-making regarding f2f classes in Fall 2020, there are a few caveats. First, our models are based on student enrollment from the 2017-2018 year, since this is the most recently available dataset. Any school that has seen a significant change (increase, decrease, or change in composition across states) in enrollment over the last two years may find that predictions are less accurate. In addition, our tool is based on IHME predictions. Thus, the accuracy of our models hinge on the accuracy of the IHME predictions. Preliminary results suggest relative agreement between our predictions and current testing results. For example, testing at West Virginia University (https://presidentgee.wvu.edu/messages/phased-return-to-morgantown-campus-july-27) suggests a ∼0.2% positive rate, which is commensurate with our estimated 0.17% positive rate. For this estimate we assumed a start date 08/30/2020, counted only active infections, and assumed an infectious period of 10 days, an incubation period of 5 days, 100% testing and a false negative rate of 0%.

Beyond the accuracy of the underlying models, our tool relies on a few additional assumptions. First, our tool allows users to pick the infectious period and incubation period. Although we preset these parameters at values consistent with CDC recommendations^10^ and existing research studies^11^, users are free to alter both, and this could impact predictions. We allow user input on these parameters because much about COVID-19 remains unknown. By allowing flexibility, our tool enables users to assess risk based on ‘best-case’ and ‘worst-case’ scenarios. In reality, both infectious period and incubation period are likely variable across the population. This brings up another assumption of our tool. Currently, our model assumes a fixed window for both of these parameters (as well as the delay between becoming infectious and dying). In a future rendition, we anticipate building in population-level variability in both infectious period and incubation period. This will improve predictions, as well as allow users to assess the importance of outliers – for instance students who shed infectious viral particles for >>10 days, or who take longer to develop an active infection after being exposed (which would require a longer quarantine to prevent detection evasion). An additional assumption of our model is that outbreaks are evenly distributed at the state level. Clearly this is not true. Indeed, some counties, even within hot-spot states, may be relatively virus-free. Students returning from these locations do not pose the same challenges as students returning from counties, cities or towns with high levels of COVID-19 infection. Currently, however, we do not have data on county-level enrollment at all of the institutes of higher education in our database. Further, IHME predictions are made at the state-level. That said, IHME anticipates building in county-level predictions in the near future, and this would facilitate increased spatial resolution in our model as well. Finally, our model does not account for international students or graduate students, as well as infections acquired during transit. Infections acquired during transit could include exposures as students travel, by car, through hot-spot cities on their way back to college. Even more likely, though, it could include exposures in airports, on buses and on trains, where there will be high levels of mixing among people from different locations across the U.S. and the world more broadly. This is almost certain to increase predicted infection rates, particularly at schools with high enrollment of out-ot-state students who are likely to return to campus by plane.

Despite the caveats, we hope that our tool proves useful in guiding decisions for the upcoming semester. Clearly, difficult choices are being made across the country as schools attempt to balance the importance of student education, financial viability, equity and access, public health and student and staff safety in the face of unprecedented challenges. Ultimately, these decisions are likely to vary based on a range of factors that determine the risks of f2f classes, as well as both the costs and benefits of remote learning. Because these decisions, and even the risks that they rest on, are not ‘one-size-fits-all’, our tool provides tailored estimates based on the incoming student body, the school location and the school capacity for mitigation via testing and quarantine.

## Data Availability

All data is either freely available on other websites, or hosted at a GitHub repository

https://github.com/bewicklab/COVID19_Consequences_of_College_Class_Continuity_Calculator

## Acknowledgements

We would like to thank Joseph Matthews, who participated in some early discussions of tool development, as well as the Clemson Covid Challenge – a summer Creative Inquiry project at Clemson University.

